# COVID-19 cases from the first local outbreak of SARS-CoV-2 B.1.1.7 variant in China presented more serious clinical features: a prospective, comparative cohort study

**DOI:** 10.1101/2021.05.04.21256655

**Authors:** Yang Song, Ziruo Ge, Shuping Cui, Di Tian, Gang Wan, Shuangli Zhu, Xianbo Wang, Yu Wang, Xiang Zhao, Pan Xiang, Yanli Xu, Tingyu Zhang, Long Liu, Gang Liu, Yanhai Wang, Jianbo Tan, Wei Zhang, Wenbo Xu, Zhihai Chen

**Author notes:** Co-corresponding authors: Zhihai Chen,; Wenbo Xu,; Wei Zhang. Yang Song, Ziruo Ge, and Shuping Cui contributed equally to this paper.

## Abstract

**Background:** The SARS-CoV-2 B.1.1.7 variant which was first identified in the United Kingdom (U.K.) has increased sharply in numbers worldwide and was reported to be more contagious. On January 17, 2021, a COVID-19 clustered outbreak caused by B.1.1.7 variant occurred in a community in Daxing District, Beijing, China. Three weeks prior, another non-variant (lineage B.1.470) COVID-19 outbreak occurred in Shunyi District, Beijing. This study aimed to investigate the clinical features of B.1.1.7 variant infection.

**Methods:** A prospective cohort study was conducted on COVID-19 cases admitted to Ditan hospital since January 2020. Data of 74 COVID-19 cases from two independent COVID-19 outbreaks in Beijing were extracted as study subjects from a Cloud Database established in Ditan hospital, which included 41 Shunyi cases (Shunyi B.1.470 group) and 33 Daxing cases (Daxing B.1.1.7 group) that have been hospitalized since December 25, 2020 and January 17, 2021, respectively. We conducted a comparison of the clinical characteristics, RT-qPCR results and genomic features between the two groups.

**Findings:** Cases from Daxing B.1.1.7 group (15 [45.5%] male; median age, 39 years [range, 30.5, 62.5]) and cases from Shunyi B.1.470 group (25 [61.0%] male; median age, 31 years [range, 27.5, 41.0]) had a statistically significant difference in median age (P =0.014). Seven clinical indicators of Daxing B.1.1.7 group were significantly higher than Shunyi B.1.470 group including patients having fever over 38°C (14/33 [46.43%] in Daxing B.1.1.7 group vs. 9/41 (21.95%) in Shunyi B.1.470 group [P = 0 .015]), C-reactive protein ([CRP, mg/L], 4.30 [2.45, 12.1] vs. 1.80, [0.85, 4.95], [P = 0.005]), Serum amyloid A ([SAA, mg/L], 21.50 [12.50, 50.70] vs. 12.00 [5.20, 26.95], [P = 0.003]), Creatine Kinase ([CK, U/L]), 110.50 [53.15,152.40] vs. 70.40 [54.35,103.05], [P = 0.040]), D-dimer ([DD, mg/L], 0.31 [0.20, 0.48] vs. 0.24 [0.17,0.31], [P = 0.038]), CD4^+^ T lymphocyte ([CD4^+^ T, mg/L], [P = 0.003]), and Ground-glass opacity (GGO) in lung (15/33 [45.45%] vs. 5/41 [12.20%], [P =0.001]). After adjusting for the age factor, B.1.1.7 variant infection was the risk factor for CRP (P = 0.045, Odds ratio [OR] 2.791, CI [1.025, 0.8610]), SAA (0.011, 5.031, [1.459, 17.354]), CK (0.034, 4.34, [0.05, 0.91]), CD4^+^ T (0.029, 3.31, [1.13, 9.71]), and GGO (0.005, 5.418, [1.656, 17.729]) of patients. The median Ct value of RT-qPCR tests of the N-gene target in the Daxing B.1.1.7 group was significantly lower than the Shunyi B.1.470 group (P=0.036). The phylogenetic analysis showed that only 2 amino acid mutations in spike protein were detected in B.1.470 strains while B.1.1.7 strains had 3 deletions and 7 mutations.

**Interpretation:** Clinical features including a more serious inflammatory response, pneumonia and a possible higher viral load were detected in the cases infected with B.1.1.7 SARS-CoV-2 variant. It could therefore be inferred that the B.1.1.7 variant may have increased pathogenicity.

**Funding:** The study was funded by the National Key Research and Development Program (grant nos.2020YFC0846200 and 2020YFC0848300) and National Natural Science Foundation of China (grant no. 82072295).

## INTRODUCTION

Since December 2019, the coronavirus disease-19 (COVID-19) pandemic caused by the highly infectious virus severe acute respiratory syndrome coronavirus-2 (SARS-CoV-2) has been rapidly spreading, which has posed a great threat to global public health^1,2^. With the continuous transmission and mutation of SARS-CoV-2, some viral variants of concern (VOC) and viriants of interest (VOI) have been reported in recent months^3-8^. On December 14, 2020, the United Kingdom reported a SARS-CoV-2 variant of concern (VOC) that belonged to the PANGO lineage B.1.1.7^9,10^, referred to as “SARS-CoV-2 VOC-202012/01”, “B.1.1.7”, 501Y.V1 or 20I/501Y.V1 (the term B.1.1.7 was used in the whole text). The B.1.1.7 variant was estimated to have emerged in late September 2020 and has increased sharply to become the predominant SARS-CoV-2 strain in England, which has soon become a global concern^7,11,12^.

With the strict prevention and control policies implemented in China, local epidemics have rarely occurred in Beijing. Occasionally, a small clustered outbreak triggered by an imported SARS-CoV-2 strain with only one chain of transmission could happen. On January 17, 2021, a clustered COVID-19 outbreak in community had taken place in Daxing District, Beijing. Confirmed by whole-genome sequencing and lineage typing results^9,10^, this outbreak was caused by SARS-CoV-2 B.1.1.7 variants. This is the first local transmission of B.1.1.7 variants in China, which constituted a new challenge to the prevention and control of COVID-19 in China. Meanwhile, three weeks prior to the Daxing outbreak, there was another local COVID-19 outbreak occurring in Shunyi district in Beijing, caused by the B.1.470 lineage which was mostly detected in Asian countries, both outbreaks had been well-controlled within a month.

Since the early transmission of COVID-19 in January, 2020, a cohort of COVID-19 cases have been established in Beijing Ditan hospital; and since June, 2020, the COVID-19 cases in Bejing have only been admitted to Ditan hospital, following which a cloud database had been established and maintained by Ditan hospital cooperating with a Beijing technology company (Beijing Zechuang Tiancheng Technology Development Co., Ltd). All relevant data for COVID-19 cases are constantly being entered into the cloud database for preparing for future prospective studies. So far, the data of about 690 cases were completely inputted. In this study, groups of two recent independent clustered outbreaks caused by distinct lineage strains and occurring in different districts in Beijing (Daxing B.1.1.7 group and Shunyi B.1.470 group) were selected as the study subjects from the cloud database. The COVID-19 cases from the two groups received alike clinical tests and treatments and each case was observed for at least 28 days. The discharge criteria included disappearance or marked improvement of clinical symptoms combined with two real-time reverse transcription quantitative PCR (RT-qPCR) tests negative on nasopharyngeal swabs more than 24 hours apart, based on *Diagnosis and Treatment Protocol for Novel Coronavirus Pneumonia Patients (Version 8)*.

Recent studies indicated that the B.1.1.7 variant has a 50% higher transmissibility^13^. Fortunately, some immunological and serological studies indicated that the B.1.1.7 variant does not escape vaccine protection^14,15^. The UK government and related scientists suggested the B.1.1.7 variant may be more deadly, but currently there is no published biological experimental evidence regarding the virulence or pathogenicity of B.1.1.7^16^.

This study is aiming at comparing the clinical presentations, the RT-qPCR results and the whole-genomic features of cases from Daxing B.1.1.7 group and Shunyi B.1.470 group, in order to evaluate the COVID-19 severity of the cases infected by the B.1.1.7 variant.

## METHODS

### Study design and recruitment of cases

This prospective cohort study were reported according to the STROBE statement and included two groups with a total of 74 confirmed COVID-19 cases from Beijing Ditan Hospital, Daxing B.1.1.7 group and Shunyi B.1.470 group, based on the different Districts and genomic typing results. The data of all the cases from the two groups were extracted from the cloud database. The confirmed cases referred to patients with clinical features of COVID-19 and positive RT-qPCR test for SARS-CoV-2 RNA at least once on the respiratory specimens collected. The Daxing B.1.1.7 group contained a total of 33 cases whereas Shunyi B.1.470 group contained 41 cases. COVID-19 patients were diagnosed according to the 8th version of *Diagnosis and Treatment Protocol for Novel Coronavirus Pneumonia Patients*, and the clinical severity was categorized into 4 grades: mild (the clinical symptoms were mild, and there was no sign of pneumonia on imaging), moderate (showing fever and respiratory symptoms, imaging manifestations of pneumonia), severe (1. dyspnea with a respiratory rate >30/min; 2. hypoxemia with oxygen saturation <93%; 3. PaO2/FiO2 <300 mmHg; 4. or the clinical symptoms worsened gradually, and the lung imaging showed that the lesions progressed more than 50% within 24-48 hours) and critical (developed complications including 1. respiratory failure that needs mechanical ventilation; 2. shock; 3. patients with other organ failure need ICU monitoring treatment). The asymptomatic cases with no clinical symptoms but positive for RT-qPCR were also required to be admitted to the hospital. Ditan Hospital is affiliated with Capital Medical University, an academic tertiary care center in Beijing, China. The hospital is also a designated center in Beijing for the diagnosis and treatment of COVID-19. Each case was observed for at least 28 days. As of this study, four cases from Daxing outbreak and one case from Shunyi outbreak were still hospitalized.

The index case of the Shunyi B.1.470 group was an asymptomatic case confirmed on December 23, 2020. He intended to leave Beijing for taking an exam, so he took the initiative to conduct nucleic acid detection before leaving in accordance with the regulations. Further epidemiological investigation indicated that he was infected by an salesperson of a mall, and the salesperson was infected by an foreign personnel who was an imported asymptomatic case that had tested negative for SARS-CoV-2 RNA within 14 days of entry. This was a clustered outbreak occurred mainly in workplaces in Shunyi District, with the majority of cases being young people. In addition, a family clustered infection including 7 members was detected.

The discovery of the index case of the Daxing B.1.1.7 group came from the routine screening of SARS-CoV-2 nucleic acid detection of a resident who intended to return home. The epidemiological investigation and nucleic acid detection of his close contacts indicated that a clustered outbreak had already occurred in the community where the case resided, of which most cases were elderly and children. Family clustered infection was mostly seen, involving a total of 13 families, of which 5 families were infected with more than 3 members and several families were infected with all members.

### Data collection

The data of the 74 COVID-19 cases from the two groups were extracted from the cloud database. Medical record review was performed to collect patients’ underlying medical conditions and symptoms at the time of diagnosis, including data of chronology of symptom onset, history of first presentation, disease progression, past medical history, physical findings, laboratory test results, imaging results, treatment and hospital course. In addition, epidemiological data of the patients were collected. Individual data were compiled into two groups of patients with the Daxing B.1.1.7 group and the Shunyi B.1.470 group for further analyses.

### Laboratory Testing

The oropharyngeal swab, nasopharyngeal swab, or sputum specimens obtained from patients during their hospital stays were collected for RT-qPCR testing. Viral RNA was extracted directly from 200-μL swab samples with a QIAamp Viral RNA Mini Kit (QIAGEN, Germany). RT-qPCR was conducted using a commercial Novel SARS-CoV-2 Nucleic Acid Test Kit (BioGerm, Shanghai, China) with a fluorescence PCR detector following the manufacturers’ instructions. A TaqMan probe–based kit was designed to detect the ORF1ab and N genes of SARS-CoV-2 in 1 reaction. Corresponding serum samples were tested for anti-SARS-CoV-2 antibodies using a chemiluminescence immunoassay (CLIA, Bioscience, Qingchong, China).

### Whole-genome sequencing and analysis

The selected swab samples of the patients were then sent to China CDC for further whole-genome sequencing. Libraries were prepared using a Nextera XT Library Prep Kit (Illumina, San Diego, CA, USA), and the resulting DNA libraries were sequenced on either a MiSeq or an iSeq platform (Illumina) using a 300-cycle reagent kit. Mapped assemblies were generated using the SARS-CoV-2 genome (accession number NC_045512) as a reference. Variant calling, genome alignment, and sequence illustrations were generated with CLCBio software. The whole-genome sequence alignment was conducted using the Muscle tool in MEGA (v7.0). Neighbor-joining phylogenetic tree was constructed using the Kimura 2-parameter model with 1,000 bootstrap replicates. Genomic lineage designation was used “PANGO lineage” typing method (https://cov-lineages.org/).

### Statistical analysis

The statistical analyses were performed using SPSS Version 24.0 (SPSS IBM, Armonk, NY, USA). We compiled data from each individual patient for the demography and clinical variables. Normal continuous variables were represented by mean and std, T-test was used to compare the statistical difference. Non-normal continuous variables were represented by median and quartile ranges, Mann-Whitney U test was used. Categorical variables were expressed as numbers and percentages, Chi-squared and Fisher’s exact tests were used to compare the statistical difference. Binary logistic regression analysis was performed to analyze the risk factors for the severity of COVID-19. All tests were two-tailed, and statistical significance was defined as P value lower than 0.05.

## RESULTS

### Whole genome sequencing and analysis of the SARS-CoV-2 samples in the two outbreaks

Clinical samples from 8 patients of the Daxing outbreak, 13 patients of the Shunyi Outbreak were sent to the China CDC for further sequencing. Some samples may be degraded due to transportation or storage. Finally, a total of 9 whole-genome sequences with good quality and coverage were obtained, including 7 from the Daxing outbreak and 2 from the Shunyi outbreak. Compared with the Wuhan reference sequence (EPI_ISL_ 402119), seven Daxing strains shared 31 nucleotide substitutions, six strains were identical, one had an addition substitution (32 substitutions) (Table 1). These seven strains shared all 28 nucleotide mutations that were first detected in the B.1.1.7 variant from the UK (Figure 2). Likewise, they had 3 amino acid deletions and 7 amino acid mutations on the spike protein corresponding to the characteristics of B.1.1.7 variants.

**Table 1.**
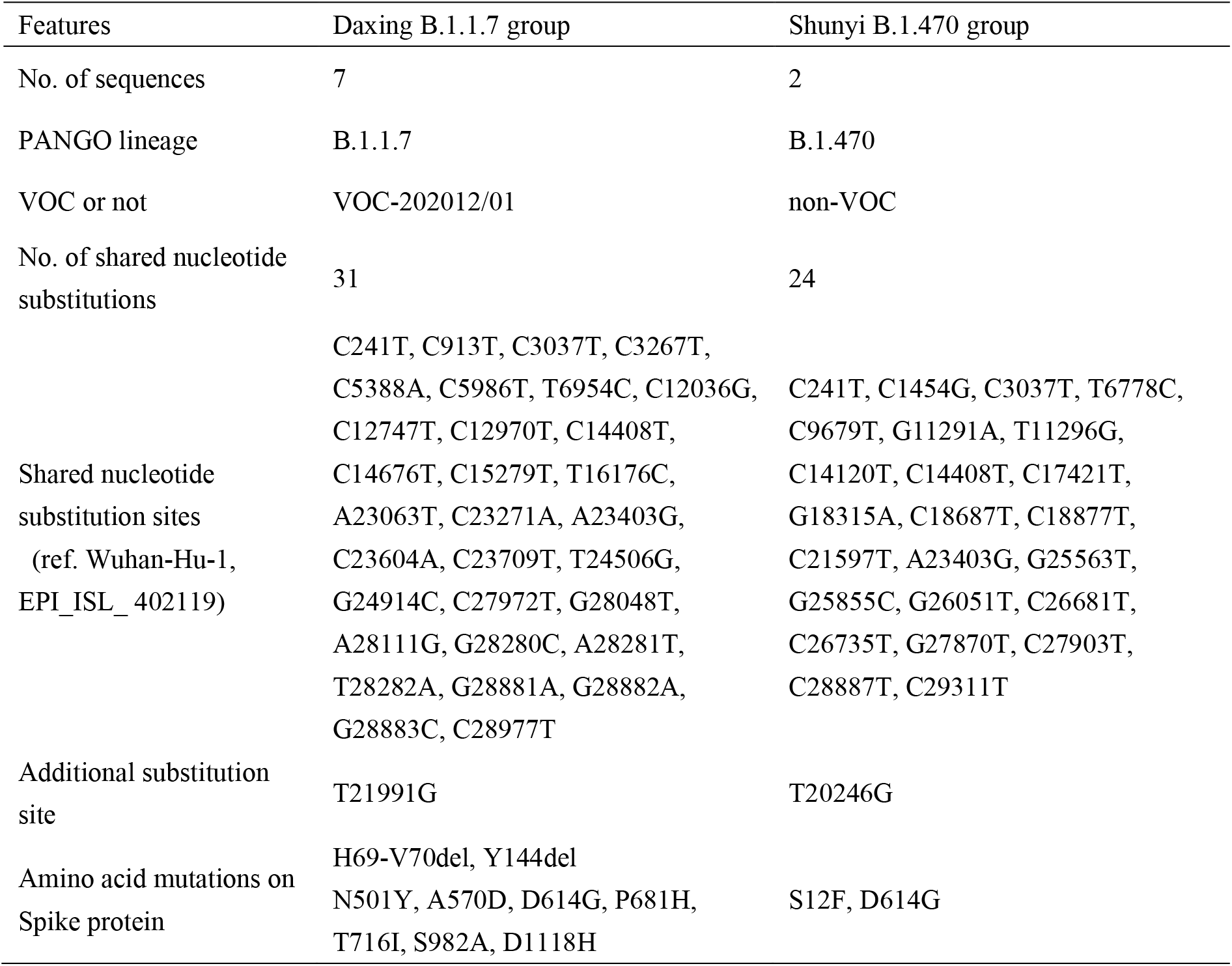
Comparison of the whole-genome features between the two groups.

| Features | Daxing B.1.1.7 group | Shunyi B.1.470 group |
| --- | --- | --- |
| No. of sequences | 7 | 2 |
| PANGO lineage | B.1.1.7 | B.1.470 |
| VOC or not | VOC-202012/01 | non-VOC |
| No. of shared nucleotide substitutions | 31 | 24 |
| Shared nucleotide substitution sites (ref. Wuhan-Hu-1, EPI_ISL_ 402119) | C241T, C913T, C3037T, C3267T, C5388A, C5986T, T6954C, C12036G, C12747T, C12970T, C14408T, C14676T, C15279T, T16176C, A23063T, C23271A, A23403G, C23604A, C23709T, T24506G, G24914C, C27972T, G28048T, A28111G, G28280C, A28281T, T28282A, G28881A, G28882A, G28883C, C28977T | C241T, C1454G, C3037T, T6778C, C9679T, G11291A, T11296G, C14120T, C14408T, C17421T, G18315A, C18687T, C18877T, C21597T, A23403G, G25563T, G25855C, G26051T, C26681T, C26735T, G27870T, C27903T, C28887T, C29311T |
| Additional substitution site | T21991G | T20246G |
| Amino acid mutations on Spike protein | H69-V70del, Y144del N501Y, A570D, D614G, P681H, T716I, S982A, D1118H | S12F, D614G |

The two strains of the Shunyi outbreak shared 24 nucleotide substitutions and one of them had an additional substitution (Table 1), which contained the single nucleotide polymorphisms (SNPs) defining PANGO lineage B.1.470 confirmed by Pangolin COVID-19 Lineage Assigner Web application^9,10^ (https://pangolin.cog-uk.io/).This lineage is composed of only a hundred more sequences on the GISIAD database, most of which (77.0%) were from Indonesia. Strains from the GISAID database^17^ which had high homology with these 2 strains were retrieved, including a Singaporean and an Indonesian strain that shared 21 and 20 nucleotide substitutions with the Shunyi strains, respectively (Figure 1).

**Figure 1.**
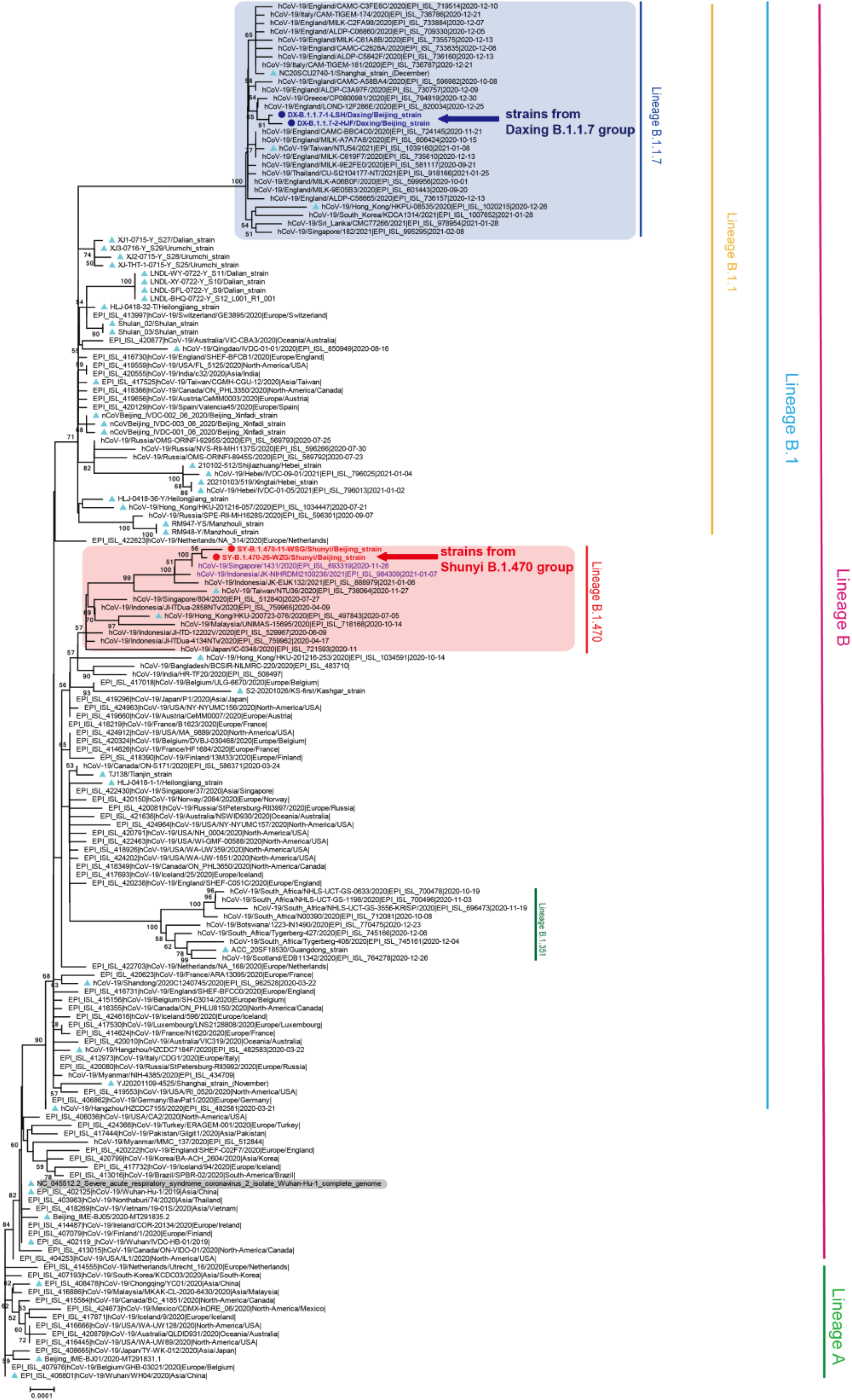
Neighbor-Joining phylogenetic tree based on the whole genome sequences of the SARS-CoV-2 representative strains. The two representative strains from the Daxing B.1.1.7 group were indicated by deep blue dots, font and a left arrow; while the strains from the Shunyi B.1.470 group were indicated by red dots, font and a left arrow. Strains associated with other previous outbreaks in China were indicated with ice blue triangles. The B.1.1.7 and B.1.470 lineage was highlighted with blue and light red background, respectively, the two strains that shared high homology with the strains of the Shunyi B.1.470 group were colored in purple font. The Wuhan reference strain was shaded in gray. The PANGO lineages were marked and colored on the right. The tree was rooted using strain WH04 (EPI_ISL_406801) in accord with the root of Pangolin tree.

**Figure 2.**
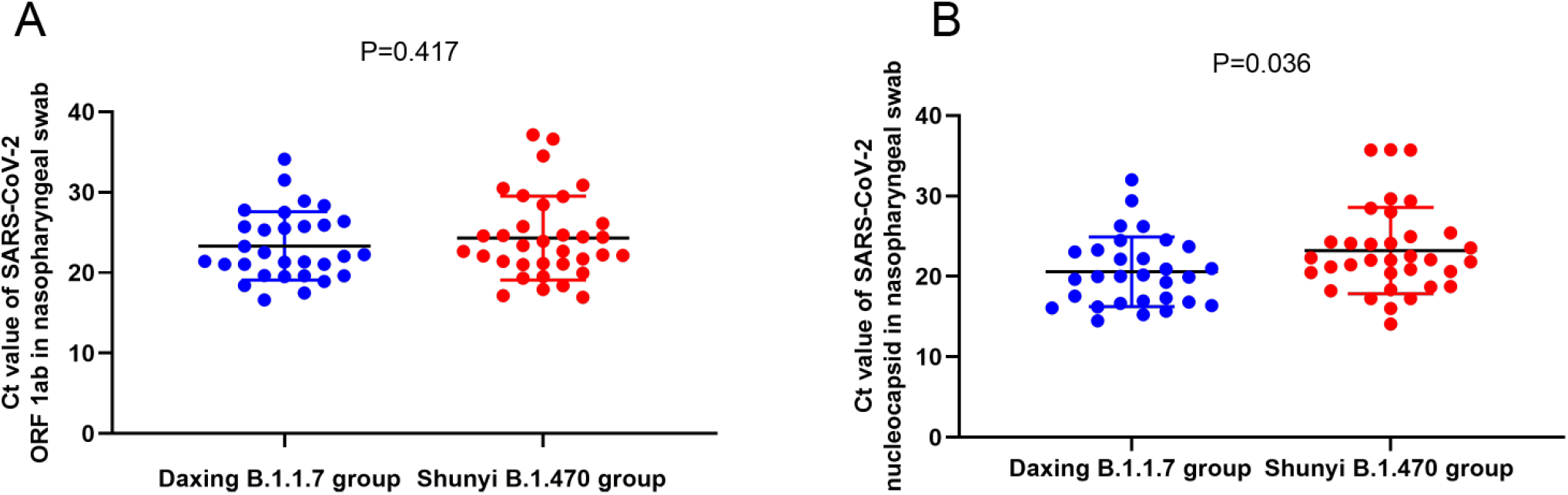
Scatter plot of the RT-qPCR Ct values of the Daxing B.1.1.7 group and the Shunyi B.1.470 group. (A) Ct values of OFR1ab-gene target; (B) Ct values of N-gene target. Samples from the Daxing B.1.1.7 group were shown as red dots while from the Shunyi B.1.470 group as blue dots. Median Ct was indicated by a black horizon bar.

### Comparison of the general information and basic clinical manifestations in the two groups

Of the 33 cases from the Daxing B.1.1.7 group and 41 from the Shunyi B.1.470 group (table1), male cases accounted for 45.50% and 61.00%, respectively. The median age of the two groups (39 years [interquartile range [IQR]30.50-62.50], vs. 31 years [IQR 27.50-41.00]) showed a statistically significant difference (P =0.014). Moderate cases took up the highest percentage of both groups (66.67% vs. 48.78%). The most common symptoms in both groups were fever, followed by dry cough and dry throat/pharyngeal discomfort. Of patients infected with B.1.1.7 variant, 14 (46.43%) had fever over 38°C, significantly higher than 9 (21.95%) observed in patients infected with B.1.470 non-variant (P =0 .015).

### Comparison of laboratory tests, imageological diagnosis and treatment measures

There were no significant differences in the levels of white blood cells, neutrophils, lymphocytes, platelets, Alanine and Aspartate aminotransferases in the two groups (Table 2). However, the level of C-reactive protein (CRP), serum amyloid A(SAA), creatine kinase (CK), and D-dimer (DD) in the Daxing B.1.1.7 group was significantly higher than the Shunyi B.1.470 group (P =0.005, 0.003, 0.040 and 0.038, respectively). In addition, except for 1 case in the Daxing B.1.1.7 group and 2 cases in the Shunyi B.1.470 group who did not tested for T lymphocyte due to their young age, the abnormal proportion of CD4^+^ T lymphocytes(CD4^+^T) in the Daxing B.1.1.7 group was significantly higher than the Shunyi B.1.470 group (P=0.003). For the imageological diagnosis, there were no significant differences in the incidence of pneumonia, but the Ground-glass opacity (GGO) observed in the B.1.1.7-variant patients was significantly higher (P =0.001).The antibody detection results and clinical treatments were not statistically different in the two groups.

**Table 2.**
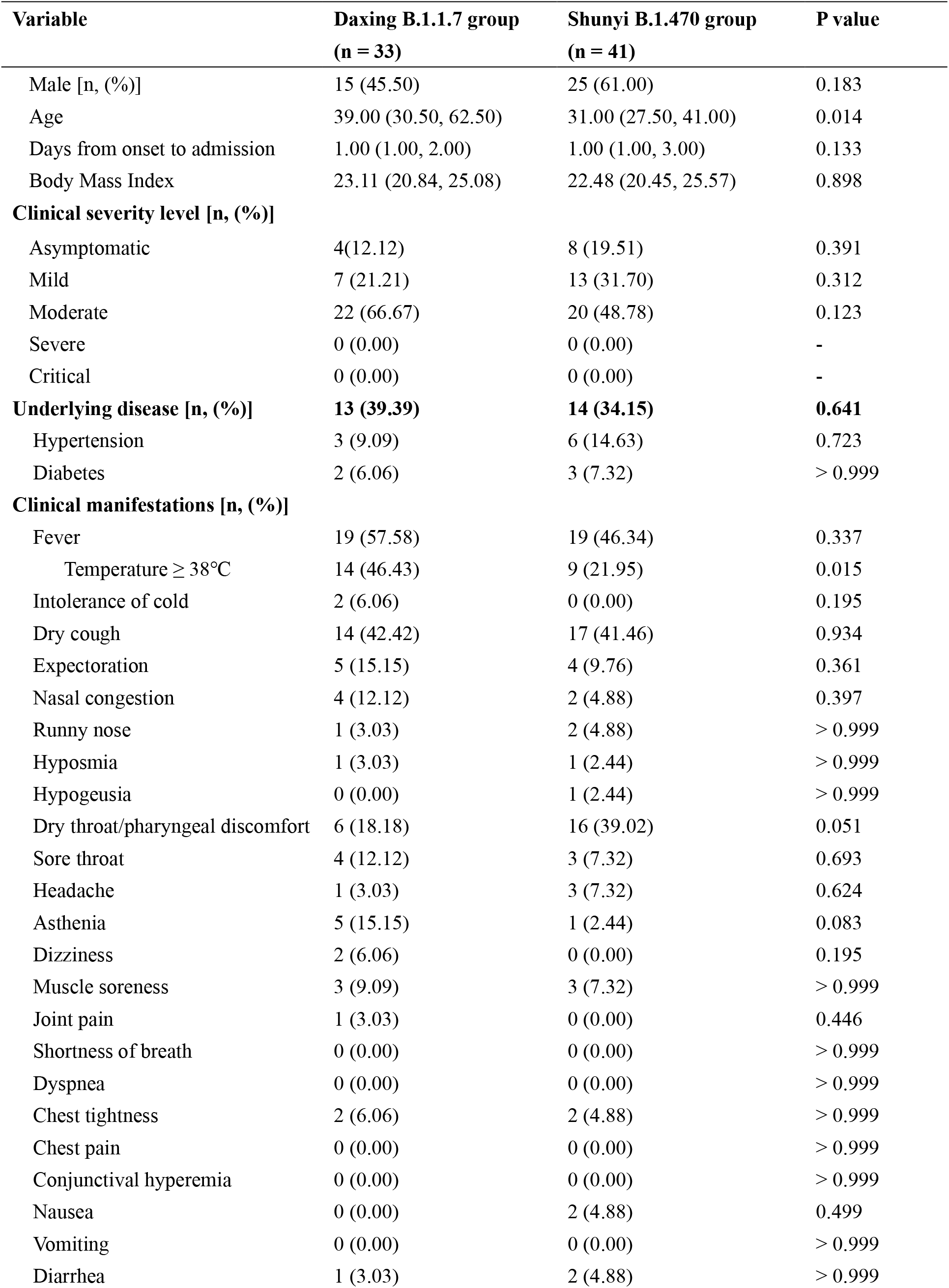
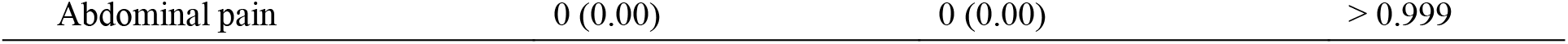
Comparison of the general situation and clinical manifestations between the two groups.

### B.1.1.7 variant infection was the main risk for more serious COVID-19 clinical features

In order to avoid the influence of older age in the Daxing B.1.1.7 group, binary logistic regression analysis was further performed for investigating the level of CRP (<7mg/L or ≥7 mg/L), SAA (<10mg/L or ≥10 mg/L), CK (<150U/L or ≥150U/L), DD (< 0.5mg/L or ≥0.5mg/L), CD4^+^ T (<706mg/L or ≥706mg/L) and GGO in the lung (Table 3). After adjusting for age factor, we found that the group factor (B.1.1.7 variant infection or non-variant infection) was the main risk for CRP (Odds ratio [OR] =2.79, P =0.045), SAA (OR =5.03, P =0.011), CK (OR = 0.22, P =0.034), GGO (OR=5.42, P=0.005), and CD4^+^ T(OR=3.31, P=0.029) of patients.

**Table 3.**
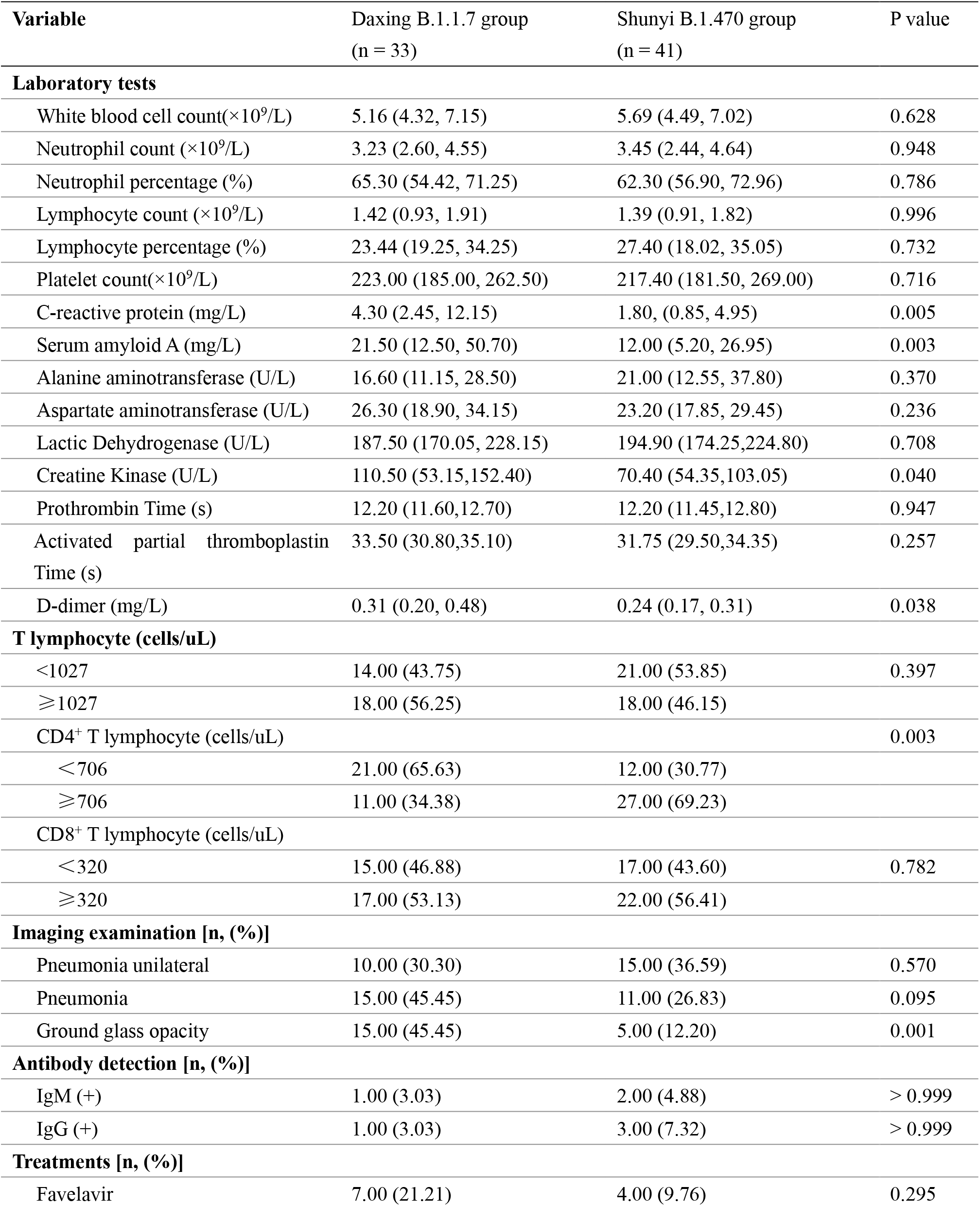
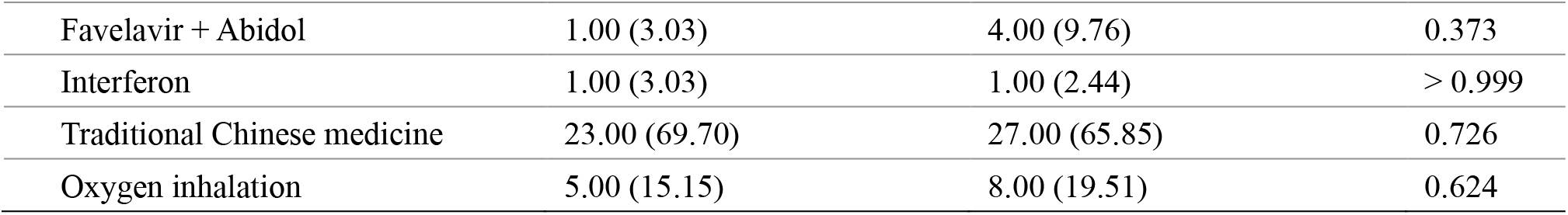
Comparison of laboratory tests, CT results and treatment measures.

### Comparison of RT-qPCR threshold-crossing (Ct) values between the two groups

During the hospitalization of all the cases, several RT-qPCR tests using nasopharyngeal swabs were regularly conducted based on their disease progression and clinical manifestations. The lowest Ct value of ORF1ab and N gene of each case was selected and compared between the two groups (Table 4). The Kits for RT-qPCR testing had uniform lot number and the instrument was the same. The median Ct value of ORF1ab-gene target in the two groups had no statistically significant difference, but that of N-gene target was significantly lower for the Daxing B.1.1.7 group than the Shunyi B.1.470 group (t=2.139, P=0.036). The distribution of the samples in the two groups was compared within all ORF1ab and N-gene Ct values (Figure 1). Figure 1b indicates that in N-gene, the median Ct values of the Daxing B.1.1.7 group were lower. In addition, we also adjusted the age factor and performed binary logistic regression analysis for the level of Ct values (>18 or ≤18) (Table 5). Similarly, patients infected with the B.1.1.7 variant had a 4.484-fold higher risk of having N gene Ct values ≤ 18 in nasopharyngeal swab specimens than the patients infected with non-variant (OR=4.484, P=0.024).

**Table 4.**
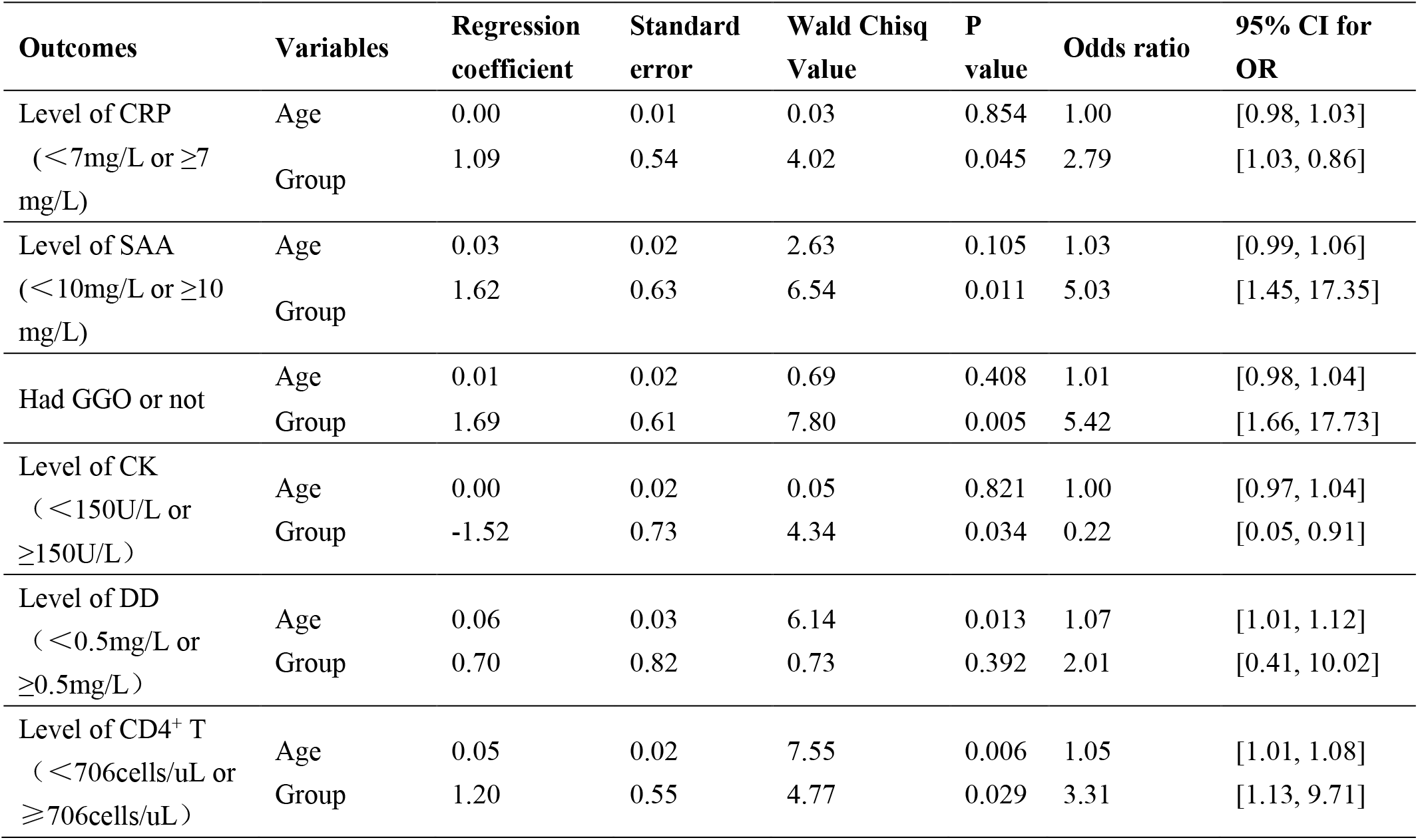
Risk of CRP, SAA and GGO of patients upon variant (B.1.1.7 variant infection or non-varient infection)

**Table 5.** Compared lowest RT-qPCR Ct values in two groups.

| Lowest Ct values | Daxing B.1.1.7 group<br>(n=33) | Shunyi B.1.470 group<br>(n=41) | P value |
| --- | --- | --- | --- |
| ORF1ab (Mean ± SD*) | 23.33 ± 4.24 | 24.31 ± 5.21 | 0.417 |
| N (Mean ± SD) | 20.58 ± 4.34 | 23.22 ± 5.37 | 0.036 |

\*SD, Standard Deviation

**Table 6.** Risk of Ct values of N gene-target of patients upon variant (B.1.1.7 variant infection or non-variant infection)

| Variables | Regression coefficient | Standard error | Wald Chisq Value | P value | Odds ratio | 95% CI for OR |
| --- | --- | --- | --- | --- | --- | --- |
| Age | -0.01 | 0.02 | 0.42 | 0.518 | 0.99 | [0.96, 1.02] |
| Group | 1.50 | 0.67 | 5.08 | 0.024 | 4.48 | [1.22, 16.53] |

## DISCUSSION

Researches have commonly indicated that the SARS-CoV-2 B.1.1.7 variant had higher transmissibility while whether it had increased pathogenicity remained controversial. In this study, we analyzed the differences of clinical characteristics, laboratory tests, RT-qPCR results and whole-genome features between the 33 cases infected with SARS-CoV-2 B.1.1.7 variant and 41 cases infected with non-variant, and indicated that COVID-19 cases infected with the B.1.1.7 variant had more serious clinical features and possibly higher viral loads. This therefore implies that the B.1.1.7 variant may have increased pathogenicity.

In the Daxing B.1.1.7 group, we observed an older age of patients than the Shunyi B.1.470 group, probably due to the living characteristics of residents from the community, as many retired older people were more frequently active in the community.

B.1.1.7 variant infection could lead to a more serious inflammatory response, acute response process and more severe pneumonia as indicated by the following results: compared B.1.1.7 variant cases with non-variant cases, (1) there were more patients with body temperature over 38°C; (2) the laboratory tests of the level of CRP, SAA, CK and DD was significantly higher; (3) the abnormal proportion of CD4^+^T was significantly higher; (4) the patients that had ground glass opacity in the lung were significantly more.

When comparing the Ct values of the RT-qPCR results between the two groups, samples from the Daxing B.1.1.7 group were related to lower Ct values of N gene, from which it could be speculated that B.1.1.7 variant samples may have a relatively higher viral load. This result corresponded with previous research showing that the B.1.1.7 variant is associated with significantly higher viral loads in samples tested by ThermoFisher TaqPath RT-qPCR^18^. Even though the sample size in this study was relatively small, and no significant difference was detected in ORF gene, statistical results for N gene could still somehow indicate a higher infectivity of the B.1.1.7 variant.

SARS-CoV-2 B.1.1.7 variant infection may be the main risk for more serious clinical features after adjusting the age factor. Old-age has been considered as an important factor in SARS-CoV-2 infection and severe COVID-19, because elderly patients have a weaker immune system function and are prone to multi-system organ dysfunction and even failure^19-22^. Thus, we adjusted the effect of older age of the Daxing B.1.1.7 group, and still came to the consistent statistically significant results that cases infected with the B.1.1.7 variant presented more serious clinical features and higher infectivity.

The phylogenetic analysis showed that the strains from the two groups belonged to different lineages. The whole genomic analysis revealed that the case samples from group-DX B.1.1.7 had 3-4 specific substitutions in addition to the 28 nucleotide substitutions corresponding to the B.1.1.7 reference sequence, which indicated that the strains might have been evolving for some time and transmitted to China. The retrieving result of the sequences that had the highest similarity with the strains of the Shunyi B.1.470 group might somehow indicate potential countries of origin. Furthermore, only two amino acid mutations in the S protein were detected in the strains from the Shunyi outbreak, which suggested lower transmissibility compared with B.1.1.7 strains.

The interaction of the SARS-CoV-2 Spike receptor binding domain (RBD) with the ACE2 receptor on host cells is vital for viral entry and the N501Y amino acid mutation of B.1.1.7 in RBD of the interface should be favorable for the interaction with ACE2^14,23,24^. Therefore, N501Y mutation likely increased transmissibility and possibly pathogenicity of B.1.1.7. Other studies indicated that the deletion of 69-70 amino acids on S1 N-terminal domain could enhance virulence and confer resistance to the neutralization antibody of SARS-CoV-2^25^. Fortunately, many recent studies on the protection of existing vaccines against B.1.1.7 variants all indicated that the B.1.1.7 variant will not escape vaccine protection^14,15,23,24^.

Nevertheless, currently there is no direct biological experimental evidence that can confirm the increased pathogenicity or virulence of the B.1.1.7 variant and little evaluation of the clinical characteristics of B.1.1.7 variant infection have been conducted. This prospective, comparative cohort study of COVID-19 cases from two COVID-19 outbreaks in Beijing implied that COVID-19 cases led by the B.1.1.7 variant presented more serious clinical and laboratory characteristics than cases infected with non-B.1.1.7 variant (lineage B.1.470).

Limitations of this study exist. Firstly, due to the very low morbidity of COVID-19 now in China, the sample size in the two groups of this study was relatively small; secondly, we were not able to compare the clinical features of the cases infected with the B.1.1.7 variant with the cases infected with other SARS-CoV-2 lineages other than B.1.470. Because during December, 2020 – January 2021, only these two clustered outbreaks had taken place in Beijing, the data of these two groups was more comparable.

## Data Availability

The data that support the findings of this study are available from the corresponding author on reasonable request. Participant data without names and identifiers may be shared with other researchers after approval from the corresponding author and the authorities including the Institutional Review Board and the National Health Commission. The proposal with a detailed description of study objectives and a statistical analysis plan will be needed for evaluation of the reasonability to request for our data. The corresponding author will make a decision based on these materials. Following complete publication, the sequence data generated in this study was made available to researchers through GISAID database (accession nos: EPI_ISL_1121993 and EPI_ISL_1122015 to 1122017). .

## Contributors

ZC conceived the study; ZC, WX and WZ designed the study; ZC and WZ maintained database for data collection; ZC, WX and WZ supervised the data collection; WZ, SC, ZG, DT, YS, GW, XW, YW, PX, YX, TZ, LL, GL and JT interpreted the data; SC, ZG and GW did the statistical analysis; YS, WZ, SC and ZG wrote the manuscript; YS and SC prepared the figures; YS, XZ, SZ and YW did the COVID-19 specimens processing and sequencing; All authors reviewed and approved the final version of the manuscript. ZC, WZ and WX are the guarantors. The corresponding author attests that all listed authors meet authorship criteria and that no others meeting the criteria have been omitted.

## Acknowledgments

We thank all physicians who participated in the management of these COVID-19 cases. The study was funded by the National Key Research and Development Program (grant nos. 2020YFC0846200 and 2020YFC0848300) and National Natural Science Foundation of China (grant no. 82072295). The funding body was not involved in the study design, clinical sample collection, data analysis, and interpretation or writing of the manuscript.

## Ethical approval

The study was approved by the Institutional Review Board of Beijing Ditan Hospital, Capital Medical University in Beijing (approval number: JDLY2020-020-01).

